# Evaluating metagenomic sequencing as a stool-based diagnostic in children with presumptive TB in Uganda

**DOI:** 10.64898/2026.01.29.26345155

**Authors:** Carolina Agudelo, Moses Nsereko, Aggrey Ainebyona, Alfred Andama, Robert Castro, Sarrah Rose Mikhail Leung, Jascent Nakafeero, Gertrude Nannyonga, Kevin Nolan, Lucas Teran, Peter Wambi, Mark G. Young, Midori Kato-Maeda, Adithya Cattamanchi, Devan Jaganath, Eric Wobudeya, Ashley R. Wolf

**Author notes:** equal contribution (Carolina Agudelo, Moses Nsereko).

## Abstract

**Background:** Stool-based molecular tests are a noninvasive option for pediatric tuberculosis (TB) diagnosis, but have lower sensitivity compared to sputum-based tests. Untargeted metagenomic sequencing (mNGS) on stool could improve sensitivity and identify new gene targets for molecular testing.

**Methods:** We performed shotgun mNGS on DNA isolated from stool samples of children undergoing assessment for pulmonary TB in Uganda. We defined the performance of mNGS to identify *Mycobacterium tuberculosis* (*Mtb*) against a microbiological reference standard (MRS, TB if sputum Xpert Ultra or culture positive) and a composite reference standard (TB if confirmed or unconfirmed TB). We also compared accuracy of mNGS against the stool-based Xpert Ultra test. Finally, we identified enriched genomic loci among *Mtb* classified reads.

**Results:** We analyzed 176 stool samples of children with a median age of 3.6 years (IQR, 1-6 years). Against the MRS, the sensitivities of mNGS with positive TB defined as ≥ 1, 2, or 5 sequence fragments were 35.5% (95% CI 19%-55%), 25.7% (12%-45%), and 19.4% (13%-25%) respectively, and specificities 92.64% (87%-96%), 97% (93%-99%), and 99.3% (96%-100%). Stool Xpert Ultra had similar sensitivity (22.6%) to stool mNGS considering all samples tested. In a head-to-head comparison, stool mNGS had lower sensitivity than stool Xpert Ultra (38.5% vs. 53.8%, difference -15.3%, 95% CI 14-68 to 25-81). mNGS utilized rRNA, virulence proteins and membrane proteins not targeted in current PCR-based platforms.

**Conclusions:** Metagenomic sequencing of stool DNA did not increase sensitivity of TB detection, but identified novel targets for molecular testing that may support development of more sensitive tests.

## Background

Tuberculosis (TB) is a life-threatening infectious disease caused by the bacillus *Mycobacterium tuberculosis* (*Mtb*) resulting in 1.3 million new cases annually in children and adolescents worldwide(1). Children have a disproportionately higher mortality risk compared to adults, in large part due to inadequate diagnostics that can delay treatment. Children often have paucibacillary disease, with a low number of bacilli in their sputum (2), which makes diagnosis more challenging. Gold-standard adult tests, including molecular PCR-based testing, typically rely on sputum, which children cannot expectorate, forcing reliance on invasive sampling of children by nasopharyngeal or gastric aspirate(3). Consequently, there is a need for novel non-sputum TB diagnostics for children.

Stool has been a promising sample type as it is non-invasive to collect and follows the same principle as gastric aspirates to detect *Mtb* from swallowed sputum. Stool-processing methods have been developed to support testing on Xpert MTB/RIF Ultra (Xpert Ultra), which performs PCR-based amplification of rifampicin resistance mutations on the gene *rpoB* and *Mtb* complex (MTBC) multicopy genes IS1081 and IS6110(4,5). Consequently, the WHO has endorsed stool-based PCR testing for pulmonary TB in children, and there have been global efforts to implement stool TB testing in routine care(6). However, meta-analyses and prospective studies have shown that stool Xpert Ultra sensitivity is lower than sputum or gastric aspirates in children (7– 11). Given the ease of stool collection compared to sputum in children, it is important to consider approaches to further optimize sensitivity for TB testing.

Metagenomic sequencing (mNGS) is an untargeted approach to broadly detect microbial DNA with high sensitivity. mNGS has been used for diagnostics, but primarily in the case of sterile samples including blood and CSF(12,13). Here we look to see whether metagenomics can be utilized in more complex and non-sterile samples that are more easily acquired. Utilizing stool samples collected during a prospective assessment of stool Xpert Ultra in children and adolescents(14), we performed mNGS to determine whether sequencing can detect rare *Mtb* reads that might not be detectable by Xpert Ultra.

## Methods

### Study Procedures

We used stool samples collected previously as described in Jaganath et al(14,15). In this study, children <15 years old were consecutively enrolled from June 2019 to March 2021 in Kampala, Uganda. Participants were recruited from inpatient and outpatient facilities at Mulago National Referral Hospital and the surrounding area. Children were eligible if they had microbiologically confirmed TB or ≥1 symptom of pulmonary TB: unexplained cough for ≥2 weeks, unexplained fever for ≥1 week, unexplained failure to thrive or weight loss, or chest radiograph suggestive of TB. Children treated for TB in the last year or on antituberculosis treatment for >72 hours were excluded. Caregivers completed a written informed consent, and children provided assent if aged 8 years or older. The study was approved by the Mulago Hospital Research and Ethics Committee, the Uganda National Council of Science and Technology, and the University of California, San Francisco, Institutional Review Board.

Study staff asked participants to collect 1 stool sample in a sterile cup, either directly or transferred from a diaper with a spoon. If the child was unable to produce a sample, a sample could be obtained at home and returned within 3 days. Stool samples were homogenized fresh or stored at 2°C–8°C and homogenized and tested within 72 hours. Three different isolation techniques were used to process stool before Xpert test Ultra (SPK, SOS, and OSF) as detailed by Jaganath et al(14). Positivity of at least one of the three isolation methods was considered to be positive for TB by Xpert Ultra (Cepheid, Sunnyvale, USA) on stool.

### TB Classification and Reference Standards

Participants were classified as having confirmed, unconfirmed, or unlikely TB based on National Institutes of Health consensus definitions(16). Confirmed TB was defined as having a positive sputum culture or Xpert Ultra (Cepheid, Sunnyvale, USA) for *Mtb*. Unconfirmed TB was defined as not having microbiological confirmation but presenting signs, symptoms and/or radiographic findings consistent with TB and when started on antituberculosis treatment responding well to treatment. Unlikely TB was defined as symptomatic at enrollment with negative microbiological testing and symptom resolution without antituberculosis treatment at the follow-up visit. A case was defined as unclassifiable if there was insufficient information or follow-up to determine TB status. Stool Xpert Ultra results were not used in the TB classification.

We used 4 reference standards. The microbiological reference standard (MRS) considered children as positive TB only if they were classified as “confirmed TB”, with both “unlikely” and “unconfirmed TB” considered as negative. The composite reference standard (CRS) included both “unconfirmed TB” and “confirmed TB” in the definition of positive for TB, with “unlikely TB” considered negative for TB. The confirmed versus unlikely reference standard eliminated the unconfirmed TB from the analysis, leaving confirmed TB as having TB and unlikely TB as not having TB. The sputum culture reference standard considered a positive culture (mycobacteria growth indicator tube, MGIT, or solid culture) as positive TB and a negative culture as negative.

### DNA isolation and shotgun whole genome sequencing library preparation

One scoop of stool was added to a sterile 2 ml screw-top tube with 500 μl 0.1 mm zirconia/silica beads, 500 µl of Buffer A (200 mM Tris-HCl, pH 8.0/200 mM NaCl/20 mM EDTA), 210 µl of 20% SDS, and 500 µl of phenol/chloroform/IAA (pH 7.9, 25:24:1). The sample was then bead beat at the highest speed for 4 minutes. The sample was spun down at 13,000 rpm and 4°C for 3 minutes. 200 µl of the aqueous layer was then passed through Monarch’s Genomic DNA Purification Kit (Cat. No./ID: T3010S/L) per manufacturer’s protocol.

Genomic DNA (gDNA) from these samples was then prepared for metagenomic sequencing using the published Hackflex protocol(17) with minor adjustments. Samples were diluted to 10 ng/µl instead of the 1 ng/µl recommended by the original protocol and a 1:10 dilution of tagmentation beads were used instead of the 1:100 dilution of the original protocol. Samples were pooled and sequenced on an Illumina NovaSeq to obtain 2 × 150 bp reads.

### Metagenomic analysis for *Mtb* reads

Samples with at least 2 million reads were analyzed. Our analysis included a range of sequencing depth from 2 million to 150 million reads. Sequences were passed through the Kraken2Uniq version 2.1.3 (18,19) pipeline where raw reads were first trimmed for quality control (QC) using Trimmomatic and host reads were removed using Bowtie2. Processed reads were then taxonomically classified using the Kraken MicrobialDB collection (August 2023 version). Reads classified by the database as *Mycobacterium tuberculosis* (*Mtb*, NCBI taxid: 1773) were then aligned using BLASTN against NCBI’s nt_prok database. Reads were considered to be true hits if they aligned to the *Mtb* genome with a percent identity (perc_identity) over 97 and a query coverage per HSP (qcov_hsp_perc) over 80. Samples were determined to be *Mtb* positive if they had greater than or equal to 1, 2, and 5 BLAST-confirmed *Mtb* sequence fragments. A set of two paired end reads was considered one fragment. In the rare cases where just one read mapped, this was still considered one fragment.

Reads classified as *Mtb* by Kraken were extracted and aligned to two *Mtb* reference genomes, H37Rv (Genome assembly ASM19595v2) and MTBC0(20), using BWA-MEM(21). The output bam files were merged using samtools(22) and visually aligned against each reference genome individually using JBrowse2(23).

### Statistical analysis

We assessed the sensitivity and specificity of stool mNGS against the MRS, CRS, confirmed versus unlikely reference standard, and a sputum culture reference standard. Samples that did not have a culture result were excluded. We also performed a concordance comparison between stool mNGS and stool Xpert Ultra; samples that did not have an Xpert Ultra result were excluded. Sensitivity and specificity analyses were completed in R version 4.2.1 using the epiR package(24).

### Data availability

Sequencing reads will be uploaded to NCBI Short Read Archive prior to publication and reads are available upon request prior to publication.

## Results

### Participant Characteristics

Our study included children with respiratory illness who were being assessed for pulmonary TB as previously described by Jaganath et al(14,15). Key demographic and sample characteristics are summarized in Table 1. The majority were under the age of 4 (n = 108 [61.4%]; median 3.6 years [IQR: 1-6]). Almost half of the children were underweight (n = 85 [48.3%]) with 11.4% of the total having severe acute malnutrition (SAM; n = 11). HIV status was available in 164 (93.2%) children. Of these, 30 (18.3%) were living with human immunodeficiency virus (HIV) and 5 (3.0%) were HIV-exposed. A total of 31 children (17.6%) had confirmed TB, with over half of the children (20 of 31 [64.5%]) being sputum Xpert Ultra positive.

**Table 1.** Sample Characteristics (N= 176)

| Characteristic | No. (%)<sup>a</sup> |
| --- | --- |
| Age, y | 3.6 (1-6) |
| < 1 | 30 (17.0) |
| 1-3 | 78 (44.3) |
| 4-10 | 56 (31.8) |
| 11-14 | 12 (6.8) |
| Male sex | 94 (53.4) |
| HIV positive (N=164) | 30 (18.3) |
| Underweight | 85 (48.3) |
| SAM | 20 (11.4) |
| Xpert positive sputum result (N=167) | 20 (12.0) |
| Sputum culture positive for *Mtb* (N=167) | 11 (6.6) |
| Xpert positive stool result (N=108) | 9 (8.3) |
| TB classification |  |
| Confirmed | 31 (17.6) |
| Unconfirmed | 92 (52.3) |
| Unlikely | 44 (25.0) |

Abbreviations: HIV, human immunodeficiency virus; SAM, severe acute malnutrition; TB, tuberculosis.
<sup>a</sup>Data represent no. (%) unless otherwise indicated (n=176 unless indicated).

We processed and sequenced 278 stool samples from these children using shotgun metagenomic sequencing (Figure 1A). After excluding samples that had a sampling depth of less than 2 million reads, the final sample size was 176. Of the 176 children included in the analysis, 31 (17.6%) had confirmed TB, 92 (52.3%) had unconfirmed TB, and 44 (25.0%) had unlikely TB.

**Figure 1.**
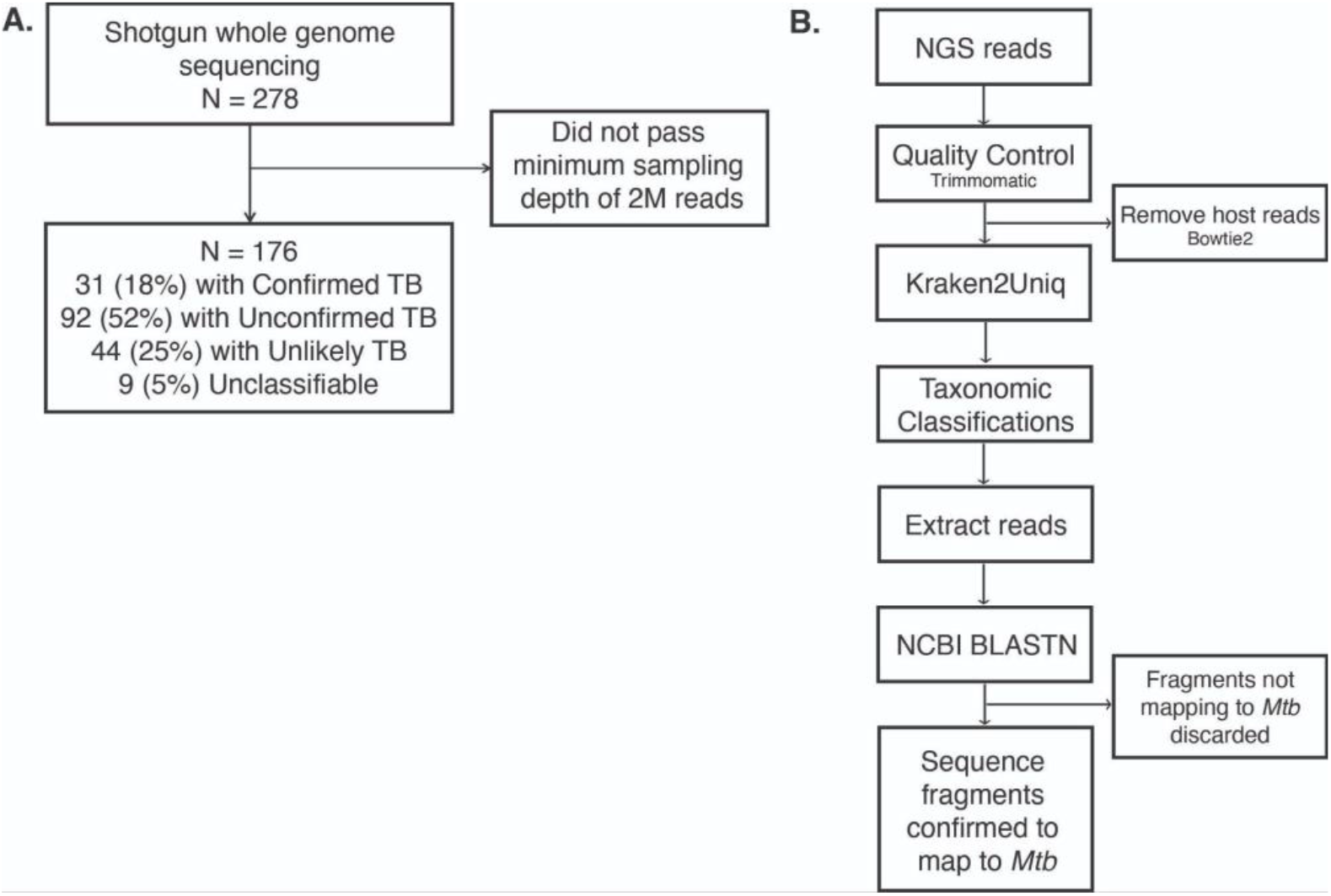
Analysis pipelines. (A) Flow chart of samples used in the analysis. (B) Bioinformatics pipeline for processing metagenomic reads for *Mtb* identification.

### Metagenomic pipeline to identify *Mtb* sequence fragments in stool metagenomes

Paired-end metagenomic reads were passed through the Kraken2Uniq(18,19) pipeline which was developed to identify pathogens in metagenomic data (Figure 1B). To verify that Kraken results were reflective of reads produced from the *Mtb* genome, we extracted reads classified as *Mtb* by Kraken and aligned them to NCBI’s nt_prok database using BLASTN. Reads were considered to be *Mtb* if they aligned to the *Mtb* genome with a percent identity (perc_identity) of over 97% and query coverage per HSP (qcov_hsp_perc) over 80%. The additional BLASTN filtering step removed 92 samples which had reads initially classified as *Mtb* but had best hits to non-*Mtb* sequences.

We also assessed whether there was a correlation between the age of the participants and how many confirmed *Mtb* sequence fragments they had as well as the sequencing depth of the samples (Supplementary Table 1). We saw an incremental increase in the average number of fragments from younger age to older children and adolescents. However, we did not see a relationship between the number of fragments and the average sequencing depth of the age group. This confirmed that there was no sequencing bias based on age.

### Accuracy of stool metagenomic sequencing to diagnose TB in children

We assessed the specificity and sensitivity of metagenomic sequencing for determining *Mtb* positivity in the cases where an *Mtb* positive was called based on greater than or equal to 1, 2, or 5 sequence fragments (Table 2). Against the MRS (where cases classified as unconfirmed TB are considered negative), the specificities of all 3 thresholds were high (92.6%-99.3%). The sensitivities at 1, 2, and 5 fragments were 35.5%, 25.8%, and 19.4% respectively. Sensitivities were lower with the CRS, which includes unconfirmed TB samples in the positive group (range, 5.69%-15.45%). When we removed unconfirmed samples and compared the confirmed TB vs. unlikely TB reference standard, sensitivities were similar to that of the MRS (range, 19.4%-35.48%). We also compared metagenomic sequencing to positivity in sputum culture (N=153). For this comparison, we removed any cultures that were contaminated or positive for a nontuberculous mycobacteria (NTM). Comparison with sputum culture positivity showed an increased sensitivity range of 45.5%-54.5%, with specificity remaining high (range, 93%-99.3%).

**Table 2.** Diagnostic Accuracy of mNGS by Read Threshold.

| Reference Standard | Sensitivity, % (95% CI) |  |  | Specificity, % (95% CI) |  |  |
| --- | --- | --- | --- | --- | --- | --- |
| | pos ≥ 1 fragment | pos ≥ 2 fragments | pos ≥ 5 fragments | pos ≥ 1 fragment | pos ≥ 2 fragments | pos ≥ 5 fragments |
| MRS | 35.5 (19-55) | 25.7 (12-45) | 19.4 (13-25) | 92.6 (87-96) | 97 (93-99) | 99.3 (96-100) |
| CRS | 15.5 (10-23) | 9 (5-15) | 5.7 (2-11) | 95 (85-99) | 98 (88-100) | 100 (92-100) |
| Confirmed vs Unlikely RS | 35.5 (19-55) | 25.7 (12-45) | 19.4 (7-37) | 95 (85-99) | 98 (88-100) | 100 (92-100) |
| Sputum culture (N=153) | 54.5 (23-83) | 54.5 (23-83) | 45.5 (17-77) | 92.3 (86-96) | 97.2 (93-99) | 99.3 (96-100) |

Abbreviations: CI, confidence interval; CRS, composite reference standard; MRS, microbiological reference standard; pos, positive; RS, reference standard.
<sup>a</sup>Data represent no. unless otherwise indicated (n=176 unless indicated).

### Comparison of metagenomic sequencing to Stool Xpert Ultra

We next compared differences in samples classified as TB positive by stool metagenomic sequencing versus Xpert Ultra on sputum or stool (Figure 2). mNGS is comparable to stool Xpert when considering the fraction of samples that are called as TB positive (Figure 2). mNGS also identified *Mtb* fragments in 8% of unconfirmed samples at the lowest threshold (pos ≥ 1 fragment), while stool Xpert Ultra only detected *Mtb* in 1% of tested samples. Sputum Xpert Ultra did not detect *Mtb* in any unconfirmed samples. Even requiring 5 fragments for positivity, mNGS detection of *Mtb* was comparable to that of stool Xpert Ultra (Figure 2).

**Figure 2.**
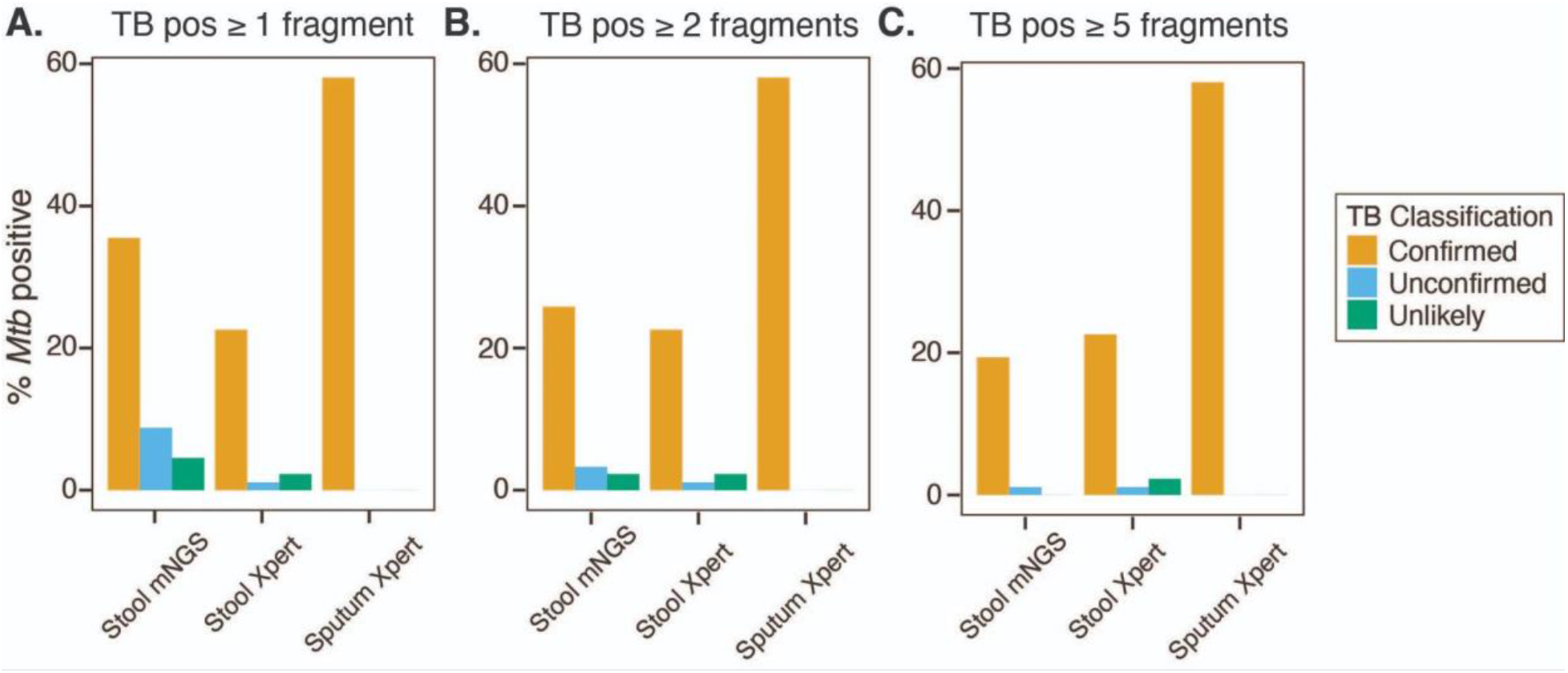
Comparison of metagenomic sequencing to Xpert Ultra on stool and sputum. TB positivity as defined by at least 1 (A), 2 (B), or 5 (C) confirmed sequence fragments mapping to *Mtb*. Percentage of samples testing positive by a given method (stool mNGS, stool Xpert, or sputum Xpert) is shown for each *Mtb* classification, confirmed (N=31), unconfirmed (N=92), and unlikely (N=44). Abbreviations: pos, positive; WGS, whole genome sequencing.

We next evaluated whether there was a correlation between the metagenomic results and sequencing depth. As mentioned previously, our analysis included a sequencing depth range of 2 million to 150 million reads. When looking at the sampling depth of the stool samples and their respective number of confirmed *Mtb* sequence fragments, we saw no relationship between the two (Supplementary Figure 1A). The samples with the highest number of *Mtb* reads had a sampling depth range of 5 to 40 million reads, indicating that metagenomic identification of *Mtb* was possible at a range of sequencing depths. We also compared the sampling depth of samples defined as positive by mNGS and Xpert Ultra on sputum and stool. Similarly, we saw no relationship between sampling depth and TB positivity in any test (Supplementary Figure 1B).

We also performed a head-to-head comparison including only samples that were tested by both mNGS and Xpert Ultra on stool (N=108, Table 3). The specificity for stool mNGS remained high when compared to Xpert Ultra stool (range 91.5%-98.9%). Stool mNGS was less sensitive when compared against stool Xpert Ultra when assessing the MRS, although the confidence intervals were large (sensitivity of 15.4%-38.5%). Considering the strictest case (> 5 *Mtb* sequence fragments), we found that only one sample was negative for both stool and sputum Xpert tests and positive by stool mNGS (Supplementary Figure 2). The remaining cases positive by mNGS were also positive for both Xpert tests or sputum Xpert sputum if stool Xpert was not tested. Comparison with a culture-based standard was not feasible as only 3 participants were culture positive with both stool Xpert Ultra and mNGS results.

**Table 3.** Head to Head comparison of stool Xpert Ultra Stool and mNGS, against the microbiological reference standard.

| Comparison | Sensitivity, % (95% CI) | Specificity, % (95% CI) |
| --- | --- | --- |
| Xpert Ultra Stool | 53.8 (25-81) | 97.8 (92-100) |
| Stool mNGS |  |  |
| pos ≥ 1 fragment | 38.5 (14-68) | 91.5 (84-96) |
| pos ≥ 2 fragments | 23.1 (5-54) | 95.8 (90-99) |
| pos ≥ 5 fragments | 15.4 (2-45) | 98.9 (94-100) |

Abbreviations: CI, confidence interval; pos, positive.

### Metagenomic sequencing captures *Mtb* genes that are not assessed by PCR-based tests

We aligned the confirmed *Mtb* aligned reads onto two *Mtb* reference genomes: H37Rv and MTBC0. When doing the first analysis with Kraken2Uniq, we saw some reads classified as other mycobacteria belonging to the *Mtb* complex (MTBC). To make sure we captured these reads, we aligned the reads to the MTBC0, an imputed ancestral reference genome(20). We also aligned the reads to the ATCC type strain, H37Rv, to represent the lineage 4 strains circulating in Uganda(25). Alignments to these genomes revealed reads aligning to multiple genomic loci (Figure 3; Supplementary Tables 2, 3). Interestingly, the genes with the most abundant gene alignments were to rRNA genes (Mtbc0_001408 and Mtbc_001409 on the MTBC0 genome; *rrs* and *rrl* on the H37Rv genome). We also looked to see if the reads aligned to genes targeted by Xpert Ultra (*rpoB*, IS6110, and IS1081). We identified fragment alignments to *rpoB*, IS6110, and IS1081 on both reference genomes (Figure 3, Supplementary Table 2, 3). Reads also aligned to the membrane protein gene *mmpL12* and several PE and PPE genes (Figure 3, Supplementary Tables 2, 3). The diversity of gene targets identified reinforces that our metagenomic taxonomic classifications are not driven by artifacts and represent true *Mtb* genome sequences. Our findings also suggest new gene targets that may be more sensitively detected than the sequences targeted by existing tests like Xpert Ultra.

**Figure 3.**
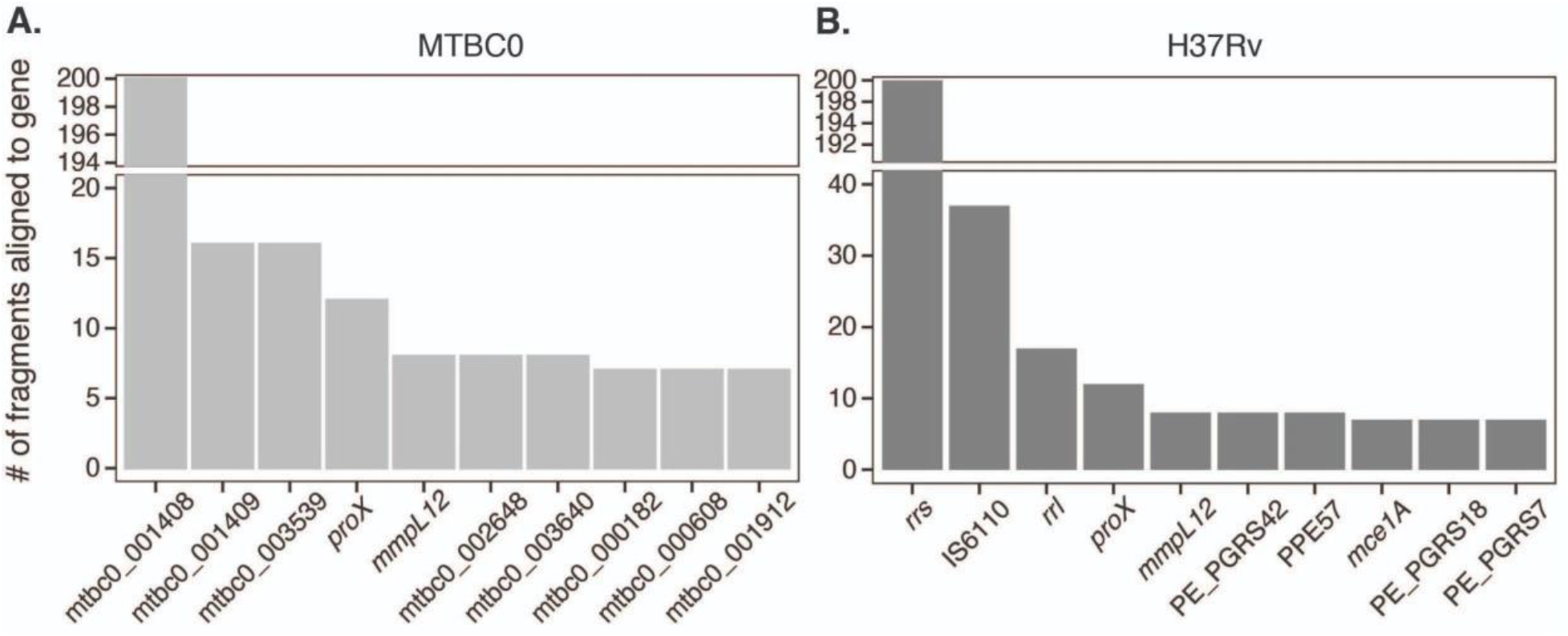
Alignment of Extracted *Mtb* Fragments Onto Reference Genomes. Top 10 sequence alignments on the (A) MTBC0 and (B) H37Rv reference genomes.

## Discussion

TB mortality rates are disproportionately higher in young children compared to adults(26,27). Young children also present unique diagnostic challenges, and as a result have poor access to accurate diagnosis and care. Diagnostic testing for pediatric TB has seen slight improvements in recent years with the advent of molecular testing on multiple specimen types(6), but sensitivity remains low.

In this study, we performed shotgun metagenomic sequencing on DNA isolated from stool of children with presumptive TB to determine if an untargeted approach like metagenomic sequencing could improve diagnostic sensitivity. We hypothesized that low sensitivity could be due to limited genomic material in the stool of some individuals or due to suboptimal selection of gene targets. We found that metagenomic sequencing was only able to capture about a third of the microbiologically confirmed cases from respiratory samples. Sensitivity almost doubled among children with culture-positive TB, suggesting that *Mtb* bacterial load in the lungs can influence the abundance of *Mtb* DNA in stool. At the same time, read alignment to reference genomes identified multicopy genes not currently targeted by Xpert Ultra that might be used to develop more sensitive tests. This overall suggests that mNGS does not add diagnostic accuracy over stool PCR, but could help identify additional PCR targets that could improve sensitivity further.

Sensitivity was low for stool mNGS regardless of the standard used, similar to what previous studies have shown for stool based PCR tests (8,14,28,29). Stool mNGS showed a comparable sensitivity to Xpert on stool for detecting culture-positive TB in children, similar to previous studies (11,14,29,30). To our knowledge, no other study has used mNGS on stool to test TB positivity. Instead, targeted next generation sequencing (tNGS) of a limited set of amplicons has been used on a variety of specimens including bronchoalveolar lavage fluid (BALF), gastric aspirate, sputum, pus, stool, and tissue(31–35). Sensitivity of tNGS in BALF and sputum tended to be higher than that of stool and tissue. Also, these studies were conducted with samples of older adolescent and adult participants. The majority of the participants of our study were under the age of 5, and on subgroup analysis this age group tended to have a lower number of confirmed *Mtb* sequence fragments (Supplementary Table 1). In recent studies, tNGS was able to detect *Mtb* DNA in 68% of PCR-positive stool DNA samples and 14-19% of stool DNA samples from individuals with paired smear-negative sputum samples(33,34). Both papers also reported sufficient coverage of antimicrobial resistance genes in the majority of the positive specimens. We detected *Mtb* DNA via mNGS in 5 of 9 Xpert stool positive cases and 12 of 162 smear-negative, *Mtb* positive cases. This suggests mNGS and tNGS may be comparable in sensitivity, but a head-to-head comparison with the same stool samples is needed to accurately define this.

These findings suggest that while the accuracy of metagenomic sequencing may be higher for culture-positive and older children and adolescents, its sensitivity is limited for infants and young children with paucibacillary TB. We found that the number of BLAST-confirmed reads was dependent on the child’s age and this was independent of sequencing depth. This suggests that enrichment for *Mtb* using tNGS rather than the deep sequencing of stool may be more promising to improve sensitivity for TB detection.

Genome alignments of metagenomic reads identified a variety of *Mtb* genes with potential as additional gene targets for current molecular testing. Currently, Xpert Ultra targets rifampicin resistance mutations on the *rpoB* gene and two multicopy genes of the MTBC, IS6110 and IS1081(4,5,36). More manufacturers have also begun to release their own assays for TB detection. For instance, Molbio released the Truenat® MTB assay which targets the *nrdB* gene and Pluslife released the MiniDock MTB Test which targets an unspecified region of MTBC DNA. However, these other assays have only been tested on nasopharyngeal and tongue swabs and sputum specimens and mostly in adult populations(37–39). Exploring relevant genes in a pediatric context may help inform manufacturer development of new and improved testing.

While our study provides insight into the poor sensitivity we see in stool testing for TB in children, it does have some limitations. The lack of a reference standard TB test for children could bias our accuracy estimates. We sought to reduce this bias by using reference standards recommended by similar research and by assessing accuracy with a range of standards and positivity thresholds. Xpert Ultra on sputum and stool was not performed for every sample that was sequenced, reducing the population for our head-to-head analyses. Due to this, we excluded several samples that were TB positive by mNGS (N=5, Supplementary Fig. 1). Future studies with additional positive samples tested by both methods are needed to confirm any variation in performance between the two methods. Whole genome sequencing also has limitations when trying to detect low abundance taxa. We identified a number of mNGS TB positives solely based on a single sequence fragment (Fig. 2), and these positives were spread across the confirmed, unconfirmed, and unlikely TB groups. These may represent a mix of false positives or true positives. Paired tNGS for these samples may provide further insight or improve sensitivity in these cases.

In conclusion, metagenomic sequencing of stool performs similarly to stool Xpert Ultra, suggesting low amounts of *Mtb* genomic material in pediatric stool. mNGS of stool may be able to capture culture negative individuals that current molecular tests miss. mNGS sequencing due to its high cost and resource requirement is not an alternative to current molecular testing for TB, but rather can provide insight into how to improve current tests. Combining mNGS with current diagnostic tests enables a comprehensive approach to understanding pediatric TB. Continued evolution and improvement of pediatric TB diagnosis is necessary to ensure children and adolescents get appropriate treatment.

## Supplementary Tables and Figures

**Supplementary Table 1.** Childhood Classification distribution of BLAST-Confirmed Reads and Sampling Depth.

| Childhood classification | N (median age) | Average number of confirmed *Mtb* sequence fragments | Average sampling depth |
| --- | --- | --- | --- |
| Infancy ($\leq 1$ year) | 30 ($\leq 1$ year) | 1.03 | 46.7M |
| Early (1-3 years) | 78 (1 year) | 0.5 | 30.3M |
| Middle (4-10 years) | 56 (6 years) | 1.38 | 33M |
| Adolescent (11-14) | 12 (13 years) | 55.75 | 32.8M |

**Supplementary Figure 1.**
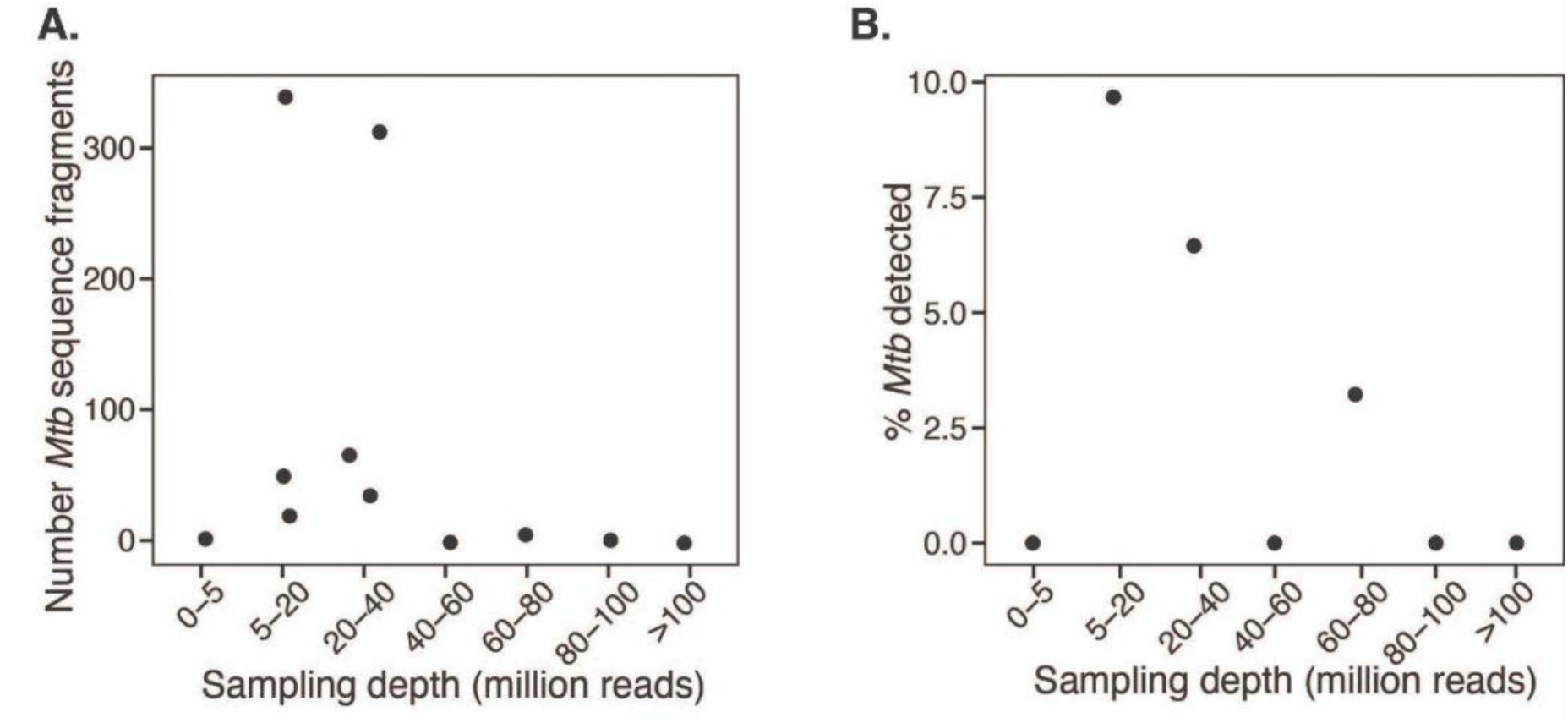
Read Depth Correlation. (A) Sampling depth of metagenomic samples and their respective number of confirmed sequence fragments (pos ≥ 5 sequences). (B) Percent of TB detected by mNGS in confirmed TB cases (N=31).

**Supplementary Figure 2.**
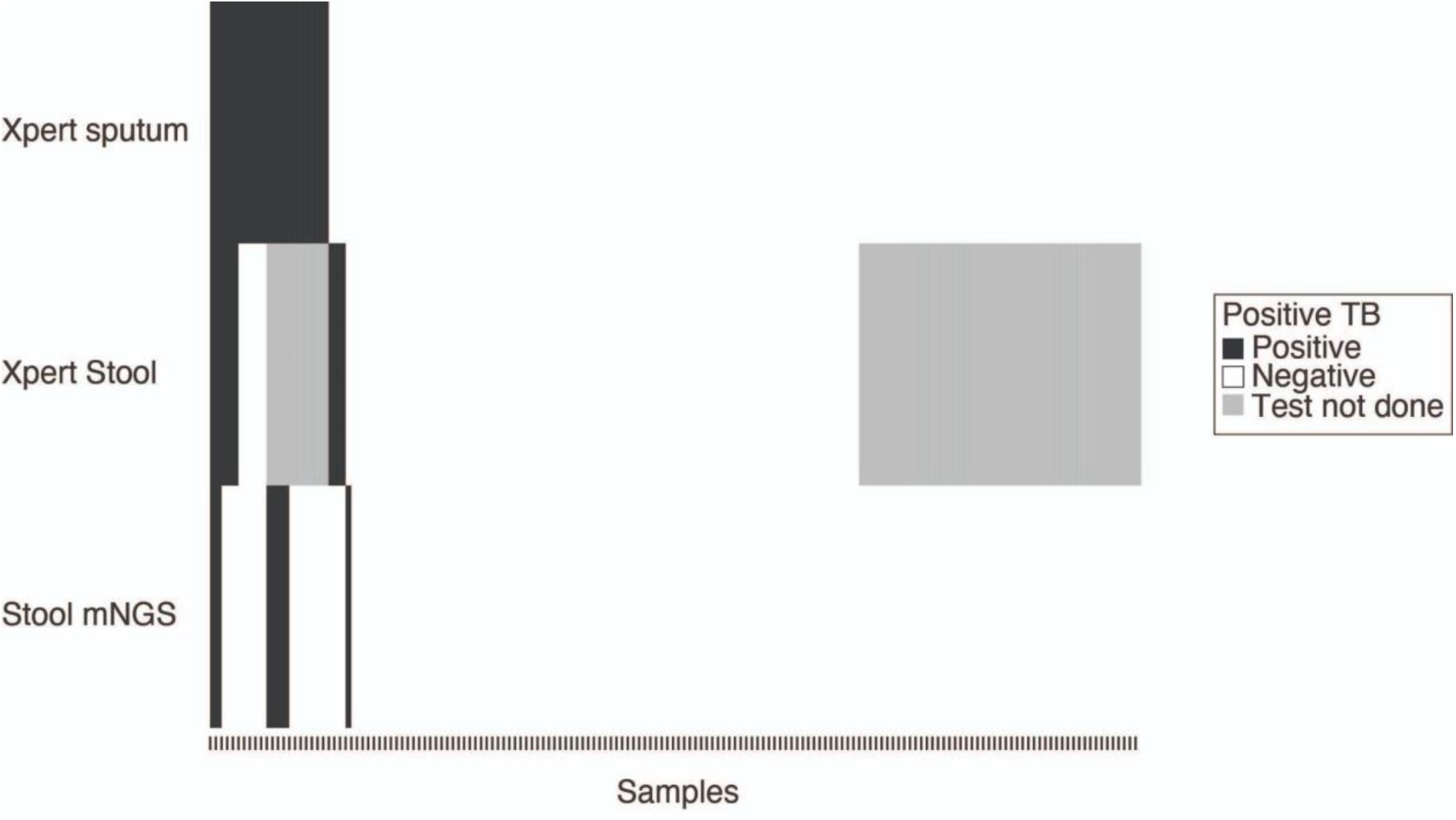
Molecular testing results on a per sample basis. Each column represents TB tests performed on the same individual. Black bars represent a positive for TB test; white bars represent a negative for TB test; gray bars denote molecular testing was not done on the sample of that participant.

